# Genomic epidemiology of SARS-CoV-2 in Pakistan

**DOI:** 10.1101/2021.06.24.21255875

**Authors:** Shuhui Song, Cuiping Li, Lu Kang, Dongmei Tian, Nazish Badar, Wentai Ma, Shilei Zhao, Xuan Jiang, Chun Wang, Yongqiao Sun, Wenjie Li, Meng Lei, Shuangli Li, Qiuhui Qi, Aamer Ikram, Muhammad Salman, Massab Umair, Huma Shireen, Fatima Batool, Bing Zhang, Hua Chen, Yungui Yang, Amir Ali Abbasi, Mingkun Li, Yongbiao Xue, Yiming Bao

**Affiliations:** China National Center for Bioinformation, Beijing 100101, China; National Genomics Data Center, Beijing Institute of Genomics, Chinese Academy of Sciences, Beijing 100101, China; CAS Key Laboratory of Genome Sciences and Information, Beijing Institute of Genomics, Chinese Academy of Sciences, Beijing 100101, China; University of Chinese Academy of Sciences, Beijing 100049, China; CAS Key Laboratory of Genomic and Precision Medicine, Beijing Institute of Genomics, Chinese Academy of Sciences, Beijing 100101, China; Department of Virology & Immunology, National Institute of Health, Islamabad 45500, Pakistan; National Center for Bioinformatics, Programme of Comparative and Evolutionary Genomics, Faculty of Biological Sciences, Quaid-i-Azam University, Islamabad 45320, Pakistan; Center for Excellence in Animal Evolution and Genetics, Chinese Academy of Sciences, Kunming, 650223, China; The State Key Laboratory of Plant Cell and Chromosome Engineering, Institute of Genetics and Developmental Biology, The Innovation Academy of Seed Design, Chinese Academy of Sciences, Beijing 100101, China

## Abstract

Pakistan has been severely affected by the COVID-19 pandemic. To investigate the initial introductions and transmissions of the SARS-CoV-2 in the country, we performed the largest genomic epidemiology study of COVID-19 in Pakistan and generated 150 complete SARS-CoV-2 genome sequences from samples collected before June 1, 2020. We identified a total of 347 variants, 29 of which were over-represented in Pakistan. Meanwhile, we found over one thousand intra-host single-nucleotide variants. Several of them occurred concurrently, indicating possible interactions among them. Some of the hypermutable positions were not observed in the polymorphism data, suggesting strong purifying selections. The genomic epidemiology revealed five distinctive spreading clusters. The largest cluster consisted of 74 viruses which were derived from different geographic locations and formed a deep hierarchical structure, indicating an extensive and persistent nation-wide transmission of the virus that was probably contributed by a signature mutation of this cluster. Twenty-eight putative international introductions were identified, several of which were consistent with the epidemiological investigations. No progenies of any of these 150 viruses have been found outside of Pakistan, most likely due to the nonphmarcological intervention to control the virus. This study has inferred the introductions and transmissions of SARS-CoV-2 in Pakistan, which could provide a guidance for an effective strategy for disease control.

## INTRODUCTION

Coronavirus disease of 2019 (COVID-19) caused by SARS-CoV-2 created a severe public health crisis, globally affecting approximately 114 million people as of Feb 28, 2021 (worldometers.info/coronavirus/). It is known that infection, transmission and death of infectious diseases are closely related to social environment such as population density, level of income and population mobility(1,2). Pakistan, as the sixth most populous country, has a population of about 223 million with limited health care resources. Since the first COVID-19 patient in Pakistan was diagnosed on February 26, 2020 in Karachi(3), over half a million Pakistanis got infected till January 2021. Although stringent measures against COVID-19 were implemented in the middle of March, the infection number kept growing and peaked in the middle of June 2020 (Supplementary Fig. 1). In Pakistan, Karachi and Lahore are the two most populous cities, while province of Punjab is most densely in population (Source: www.Worldometers.info, Supplementary Table 1). Besides, as a Muslim country (about 96.4% Muslim), religious gathering activities are frequent, which probably contribute to the spread of the disease. Fortunately, the newly diagnosed cases began to decline in late June, which suggested that novel control measures in Pakistan have played a positive and effective role. Currently, Pakistan is experiencing the second wave of infection with around 1000 new cases diagnosed per day.

**Fig. 1.**
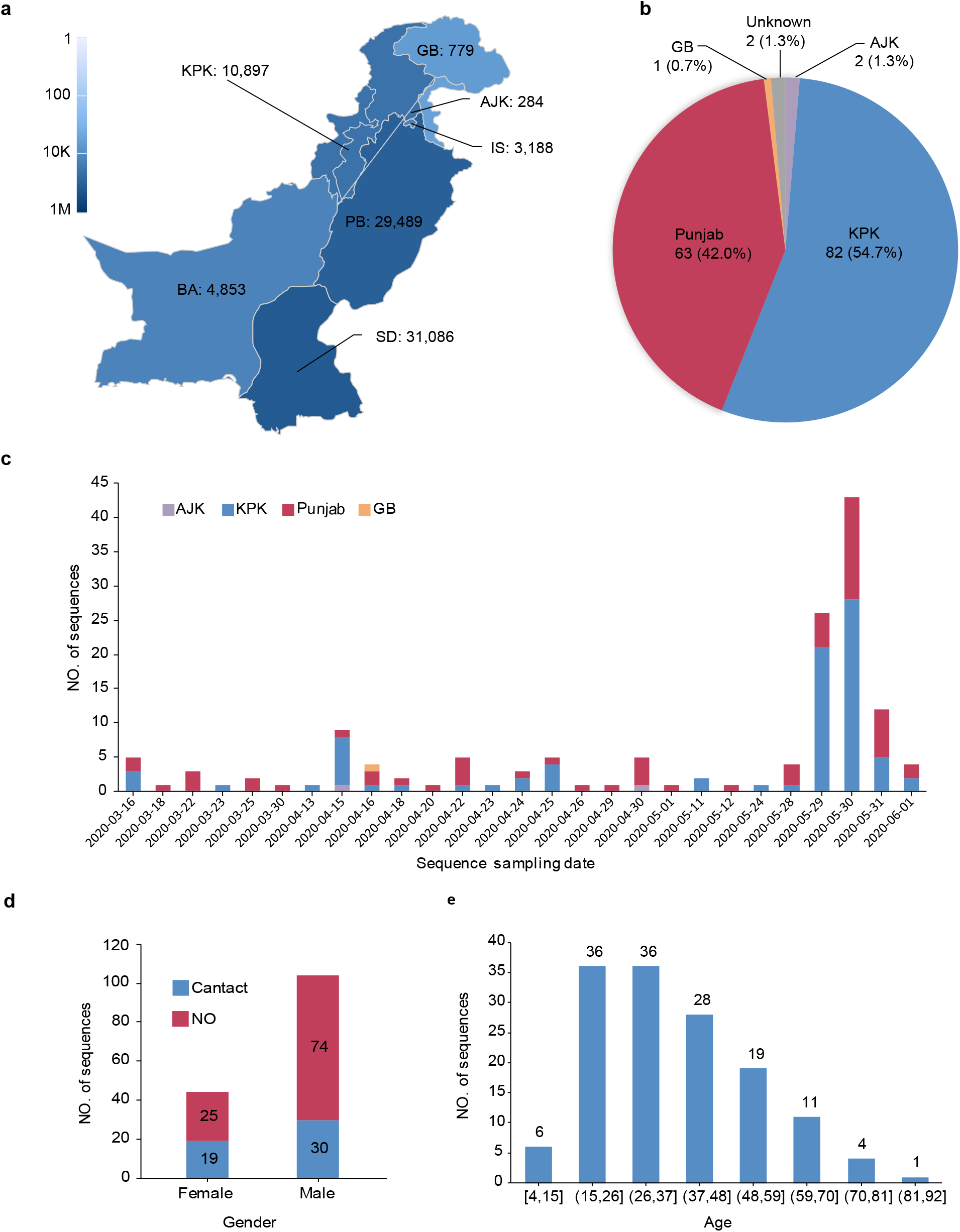
Epidemic in Pakistan and demographic details of the sampling subjects. **a** Map of Pakistan district shaded by the incidence of confirmed cases numbers as of Jun 2, 2020 as defined by the color-bar. **b** Distribution of collected samples numbers in different district. **c** Sample number on each sampling day and in each administrative region. **d** The statistic number of samples according to sex and contact history. **e** The statistic number of samples in different age groups.

Where the virus was introduced and how it was transmitted in Pakistan is largely unknown. Genomic epidemiology is a powerful tool to answer these questions. However, to date, only few dozens of viral genomes from Pakistan were publicly released(4-8). To better understand the introductions and transmissions of SARS-CoV-2 in Pakistan, we sequenced oropharyngeal samples collected during March to June from 150 COVID-19 patients, with the viral nucleic acids enriched by a hybridization capture method. We further investigated the connections between the Pakistani sequences and the potential source of acquisition by putting the high-quality viral genomes into the context of the global diversity of SARS-CoV-2 genomes. These data and the genomic epidemiology analysis allowed us to understand the transmission dynamics and contribute to more targeted and effective response to the pandemic in Pakistan. Especially, it would aid in emphasizing the measures taken by the health sectors in Pakistan and worldwide to abate the risk of further spreading of the pandemic.

## MATERIAL AND METHODS

### Case definition and sample collection

Suspected cases for COVID-19 were defined with the following clinical symptoms: acute onset of fever, cough; general weakness/fatigue, headache, myalgia, sore throat, coryza, dyspnoea, anorexia/nausea/vomiting, diarrhoea, or altered mental status, and in epidemiological working or travelling from an area with high risk of transmission of virus or health care workers. Coordinated activities in collaboration with all provinces of Pakistan were carried out at the National Influenza Center (NIC) located in NIH, Islamabad. Samples were collected from sentinel sites, secondary and tertiary care hospitals, and high risk area of COVID 19 transmission. Predesigned form were used to collect patient demographic, clinical history and risk factors according to WHO clinical and epidemiological suspected case definition(9). Samples from case contacts of patients were also collected and processed.

### Laboratory diagnosis

Throat and/or nasopharyngeal swabs were collected from suspected cases in 2-3 ml of viral transport medium (Virocult®) and stored at -80°C. RNA was extracted using Qiagen QIAmp Viral RNA mini kit and eluted in 60μl of elution buffer, and analyzed by one-step real-time reverse transcription-polymerase chain reaction (rRT-PCR) on Applied Biosystems platform ABI 7500 following the WHO Real-Time RT-PCR recommended protocol(10). The assay was performed using AgPath-IDTM One-Step RT-PCR (Ambion; California, USA). Briefly, 25μl of PCR mixture containing 0.5μl each of probe (FAM labeled), forward and reverse primers, 1μl of enzyme mix, 12.5 μl of 2X master mix, 5 μl of nuclease-free water and 5μl of extracted RNA was subjected for amplification with the following conditions: reverse transcription at 55°C for 10 min, Taq inhibitor activation 95°C for 3 min, and 45 cycles of 95°C for 15s and58°C for 30s. Inactivated RNA samples on dry ice were sent to China National Center for Bioinformation (CNCB) by air.

### Metatranscriptome library preparation

The concentration of SARS-CoV-2 was quantified with RT-qPCR targeting the *orf1ab* gene (Real-Time Fluorescent RT-PCR Kit for Detecting 2019-nCoV, BGI). Total nucleic acid was concentrated using RNA Clean & Concentrator™-5 Kit (Zymo Research) based on the manufacturer supplied protocol. The concentrated samples were converted into Illumina-compatible sequencing libraries with Trio RNA-Seq Kit (Nugen). First, DNA leftover was digested through DNase treatment. After first-strand cDNA was synthesized, 8 μl of cDNA was used for multiplex PCR sequencing, and 10μl of H2O was added to the remained 15μl product for the second strand cDNA synthesis. SPIA amplification was applied to amplify cDNA. Resultant DNA was subjected to fragmentation, end repair, and adaptor ligation through 6 cycles of PCR amplification. The DNA fragments bound to AnyDeplete probes were discarded to reduce subsequent amplification of human ribosomal RNA. Finally, eight cycles of PCR were applied, and the product was purified with AMPure XP beads (Beckman).

### Hybrid capture-based enrichment and sequencing

A 750 ng input from the library was used for hybrid capture-based enrichment of SARS-CoV-2 with two rounds of hybrid capture-based hybridization (TargetSeq® One nCov Kit, iGeneTech). Each enrichment was carried out based on manufacturer’s instructions using a hybridization temperature of 65°C for 16 hours and followed by 22 cycles of PCR in the first round, and 14 cycles in the second. Sequencing was performed on Illumina HiSeq X Ten platform with 32 samples multiplexed in a lane.

### Sequenced reads preprocessing and variants calling

Sequenced reads were preprocessed by eliminating low-quality bases (Q20 < 10), adapter sequences, and reads with length < 30 bases using Cutadapt(11). High-quality reads were mapped to the reference genome of SARS-CoV-2 (GenBank: <u>MN908947.3</u>)(12) using Burrows-Wheeler Aligner (BWA)(13). Reads with high mapping quality (MQ > 25) were retained by SAMtools(14), and duplicated reads were marked with MarkDuplicates package in Genome Analysis Toolkit (GATK)(15). Genomic variants were identified using uniquely mapped reads by HaplotypeCaller package in GATK. The genotype was assigned as a mutant allele if the frequency of mutant allele is >= 0.7, and a degenerate nucleotide was assigned if the frequency of mutant allele was <0.7 and >=0.3. The reference allele was assigned otherwise. Variants (MAF > 0.3) effects on protein-coding was annotated by VEP (Ensembl Variant Effect Predictor)(16). The depth of sequencing and the coverage of genome was calculated based on those high quality mapped (HQM) reads without duplications.

### Detection of intra-host variations

To identify the intra-host variant, mpileup files were generated by samtools v1.8 and then parsed by VarScan v2.3.9 along with an in-house script to identify intra-host variants. All intra-host variants identified had to satisfy the following criteria: (1) sequencing depth ≥ 100, (2) minor allele frequency ≥ 5%, (3) minor allele frequency ≥ 2% on each strand, (4) minor allele counts ≥ 10 on each strand, (5) strand bias of the minor allele < 10, (6) minor allele was supported by the inner part of the read (excluding 10 base pairs on each end), and (7) minor allele was supported by ≥ 10 reads that mapped exclusively to the genome of Betacoronavirus by Kraken v2.0.8-beta on each strand.

### Substitution rate estimation

To estimate the substitute rate at each position, SARS-CoV-2 genome variations and corresponding metadata were downloaded from 2019nCoVR (bigd.big.ac.cn/ncov/variation)(6,8). Phylogeny and ancestral sequence were constructed by IQtree (version 2.1.1). MN908947.3 was used as the outgroup of the phylogenetic tree. Substitution on each branch was inferred by comparing to its parental node. The substitution rate of each position was represented by the sum of the substitution events observed on the phylogenetic tree.

### Inference of the concurrency of iSNVs

The squared coefficient of correlation (r^2^) was calculated for each verified iSNVs pairs (satisfying aforementioned criteria). Position pairs with r^2^ higher than 0.8 indicated that these mutations tended to occur simultaneously. Wilcoxon signed-rank test were then performed to compare MAF at these two positions.

### Genome assembly construction

The consensus sequence for each sample was built based on the reference sequences and filtered variants using in-house Perl scripts. Specifically, the bases of mutation sites in the reference genome were replaced with their altered alleles. When sequence variation is heterozygous, the base was marked as a degenerate base symbol. And, if the mapping depth of a base is lowered than 30, it was considered to be an unknown base and marked as N.

### Haplotype network construction and phylogenetic analysis

Global sequences including 14 Pakistani sequences were downloaded from 2019nCoVR database (https://bigd.big.ac.cn/ncov) in CNCB(6,8) and GISAID EpiCoV database(17,18) as of Oct. 9, 2020. Among them, 54204 sequences sampled before Jun 17, 2020 were used for further analysis. Along with the 150 Pakistani SARS-CoV-2 sequences from this study, haplotype network was constructed as in the 2019nCoVR(8). Briefly, short pseudo sequences that consist of all variants (excluding those in the UTRs) were generated for all 54354 sequences. All these pseudo sequences were clustered into groups, and each group (a haplotype) represents a unique sequence pattern. The haplotype network was inferred from all identified haplotypes, where the reference sequence haplotype was set as the starting node, and its relationship with other haplotypes was determined according to the inheritance of mutations. JSP (Java Server Pages), HTML and D3.js (a JavaScript library for manipulating documents based on data; https://d3js.org) were employed to visualize the haplotype network. In order to show the transmission of SARS-CoV-2 in Pakistan more clearly, the sub-network directly related to Pakistani sequence were constructed. Based on the network topology, clusters were defined. The lineage for each sequence was also determined by Pangolin (version v2.0.8, Phylogenetic Assignment of Named Global Outbreak LINeages and pangoLEARN version 2020-02-18)(19)

To construct a phylogenetic tree, 164 Pakistan SARS-CoV-2 sequences and those closely related public sequences, and lineage representative sequences were used. Multiple sequence alignment was performed with MUSCLE v 3.8.31(20), and the UTR sequences of all sequences were truncated based on nucleotide coordinates of the reference genome (GenBank: MN908947.3)(12). A Maximum Likelihood phylogeny was then inferred under the best-fit model of nucleotide substitution ‘GTR+F+I+G4’ in IQ-TREE(21,22), and tree topology was assessed with the fast bootstrapping function with 1000 replicates.

### Introductions inferences

Introduction events were counted as the most recent common ancestor nodes of all 150 Pakistani sequences, plus those that had foreign travel history.

## RESULTS

### Demographic details of the subjects

From the distribution of confirmed cases (as of June 2 2020) among administrative regions in Pakistan (Fig. 1a), Sindh, Punjab and Khyber Pakhtunkhwa (KPK) have the largest number of infected individuals, with exceeding 10 thousand cases. Among them, 31,086 (39%) and 29,489 (37%) cases were from Sindh and Punjab which borders on India, and 10,897 (14%), 4853 (6%), and 779 (1%) were from KPK, Balochistan, and Gilgit Baltistan which borders on Afghanistan, Iran and China. To improve our understanding of the introductions and transmissions of SARS-CoV-2 in Pakistan, oropharyngeal swab samples were collected from 150 COVID-19 patients in Pakistan from March 16 to June 1, 2020 by the National Institute of Health in Pakistan. Most samples were collected from KPK (55%) and Punjab (42%), the two provinces with severe epidemic (Fig. 1b), while others were from Azad Kashmir (2), Gilgit Baltistan (1), and unknown regions (2). Approximately half samples were collected on May 29-30 (Fig. 1c). Based on the epidemiological investigation, the male-to-female gender ratio is 104:45 (gender information is missing for one patient), and one third of cases had contact history (Fig. 1d). The mean age was 37.6 ± 16.1, which ranged from 4 to 85 years (Fig. 1e). In all, the gender and age distribution was proportional to the affected early cases in Pakistan(3). Notably, two patients had travel history, one (E120) from Sweden and the other (E136) from Qatar. The detailed clinical information for all samples is shown in Supplementary Table 2.

### Genome sequences and variants analysis revealed Pakistan over-represented mutations

For all samples, we tested the Ct value of quantitative reverse transcription PCR targeting SARS-CoV-2 and found it ranged from 16 to below detection limit, with a median of 24. Nucleic acids of SARS-CoV-2 were enriched by the probe hybridization method and sequenced on Illumina Xten platforms. On average, 12 million paired-end reads were obtained for each sample, and a median number of 228,310 (range 5,163-358,458) reads per million (RPM) reads could be mapped to the SARS-CoV-2 reference genome (GenBank: MN908947.3). The median sequencing depth was 118,779 (ranged from 4,896 to 769,788), and 94% of the samples (141 of 150) had more than 95% of the viral genome covered by at least 100-fold (Supplementary Fig 2a, Supplementary Table 2). Based on Spearman correlation analysis, both the number of sequenced SARS-CoV-2 reads and the genome coverage were negatively correlated with the Ct value (Spearman’s correlation coefficient *r* = -0.45 and 0.61, *p* < 0.01, Supplementary Fig. 2b).

**Fig. 2.**
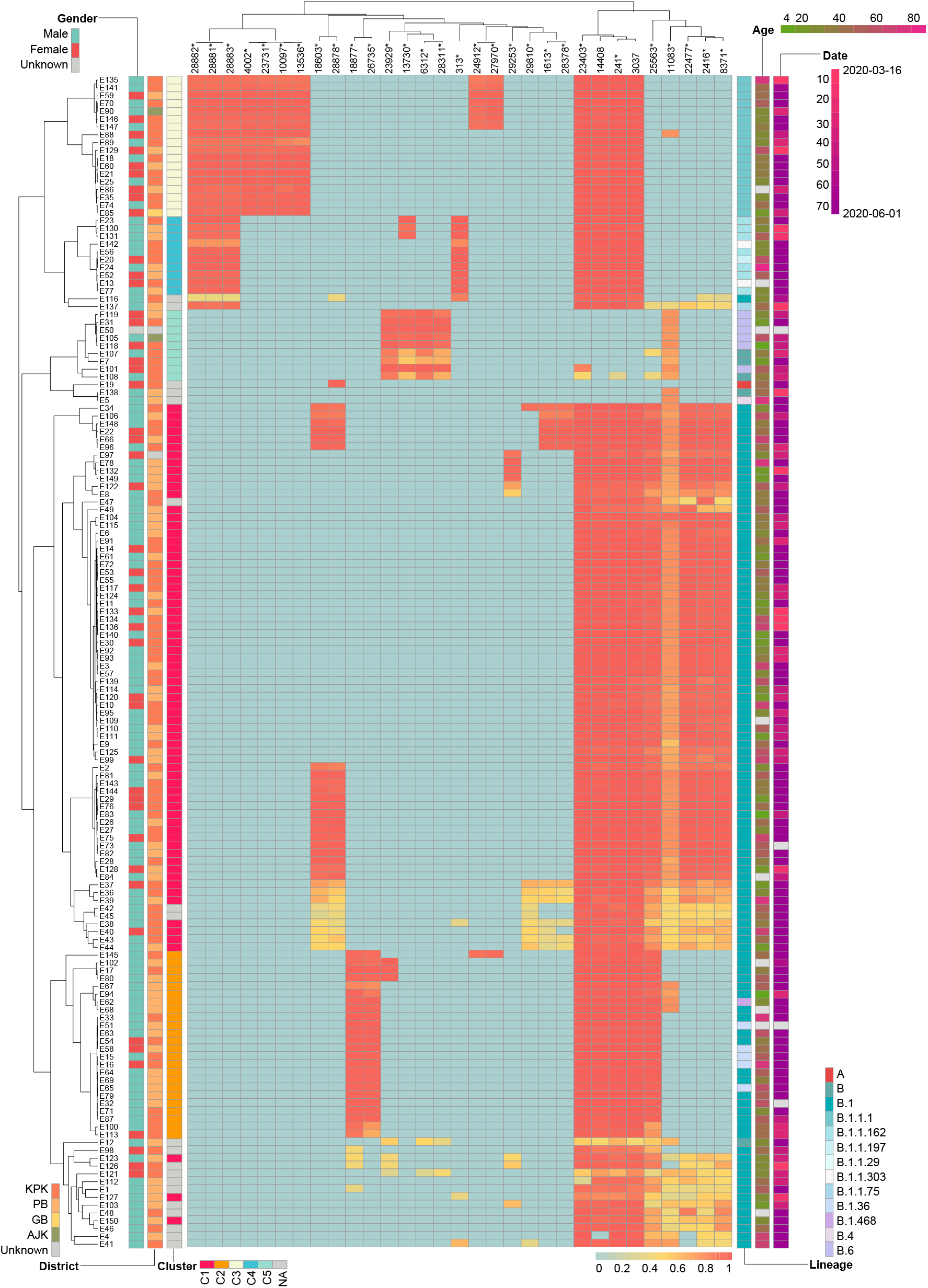
Heatmap of the mutated allele frequency (MAF) of variants with population mutated frequency (PMF) >0.05 in each sample. The lineage, and cluster information for each sample were also shown on the right.

Sequence variants for each sample were identified by aligning sequenced reads to the reference genome (GenBank: MN908947.3). The number of variants (with mutated allele frequency, MAF > 0.3) ranged from 5 to 23, with a median number of 12, which was slightly higher compared with those of global public sequences (Supplementary Fig. 2c). In total, 347 mutation sites were identified, with 103 (30%) and 171 (49%) were synonymous and nonsynonymous changes, respectively (Supplementary Table 3). By calculating the population mutated frequency (PMF) for each variant, we found 31 variants with PMF > 0.05 (Fig. 2), which means they appeared in at least 8 samples. Among them, 29 were over-represented (*p*< 0.05, Fisher’s exact test) in Pakistani sequences when compared to the PMFs in all public sequences (as of Oct. 9 2020). Consistent with the global trend(23,24), a high frequency of the viruses (140, 93.33%) harbored the D614G amino acid mutation in the Spike protein, conferred by a SNP at nucleotide 23403. Besides, more than 20 Pakistani specific mutation sites were detected, but with very low population incidence. According to the MAF of variants (PMF > 0.05) in each sample, we also observed some variants have low MAF in multiple samples, such as variants at position 2416, 8371, 11083 (Fig. 2), which suggested that there are some intra-host single nucleotide variants (iSNVs).

### Intra-host single-nucleotide variants and linkage disequilibrium between different loci

To further investigate the characteristic of iSNVs, we identified 1057 iSNVs at 513 genomic locations (MAF > 0.05), including 599 non-synonymous, 401 synonymous, 8 stop-gain, 1 stop-loss, and 48 intergenic mutations. The number of iSNV per samples was unevenly distributed, which ranged from 1 to 109, with a median of 3 (Fig. 3a). The number of iSNVs was negatively correlated with MAF, except for MAF 0.8-0.95, which may reflect back mutations (Fig. 3b). We noted that 69.7% of the mutations (737/1057) were polymorphic in the population, and the iSNVs with higher MAF were more likely to be fixed in the population (*p* < 0.01, Fisher’s exact test). The overall Ka/Ks ratio was 0.36, indicating a purifying selection on the viral genome (*p* < 0.001), which is also true for individual genes including *orf1a, orf1b, S*, and *M*. Notably, all these genes showed similar pattern in the population data (Fig. 3b). *orf3a* showed a significant signature of positive selection with Ka/Ka=6.83, which was mostly (57/76) attributed to the non-synonymous mutations occurred at two hypermutable positions in this gene, namely 25406 and 25563. Although position 25406 was prone to mutating in the individual, it had not been observed as a substitution in the polymorphism data (Fig. 3c), suggesting that the mutant was unlikely to be favored by natural selection. In contrast, G to T mutation at position 25563 defined a major branch (GH) and occurred 54 times on the phylogenetic tree of SARS-CoV-2 genomes, and its substitution rate was ranked 8^th^ among all genomic positions. However, it was still insufficient to infer a positive selection at this position, thus more analysis and especially experimental validation is needed.

**Fig. 3.**
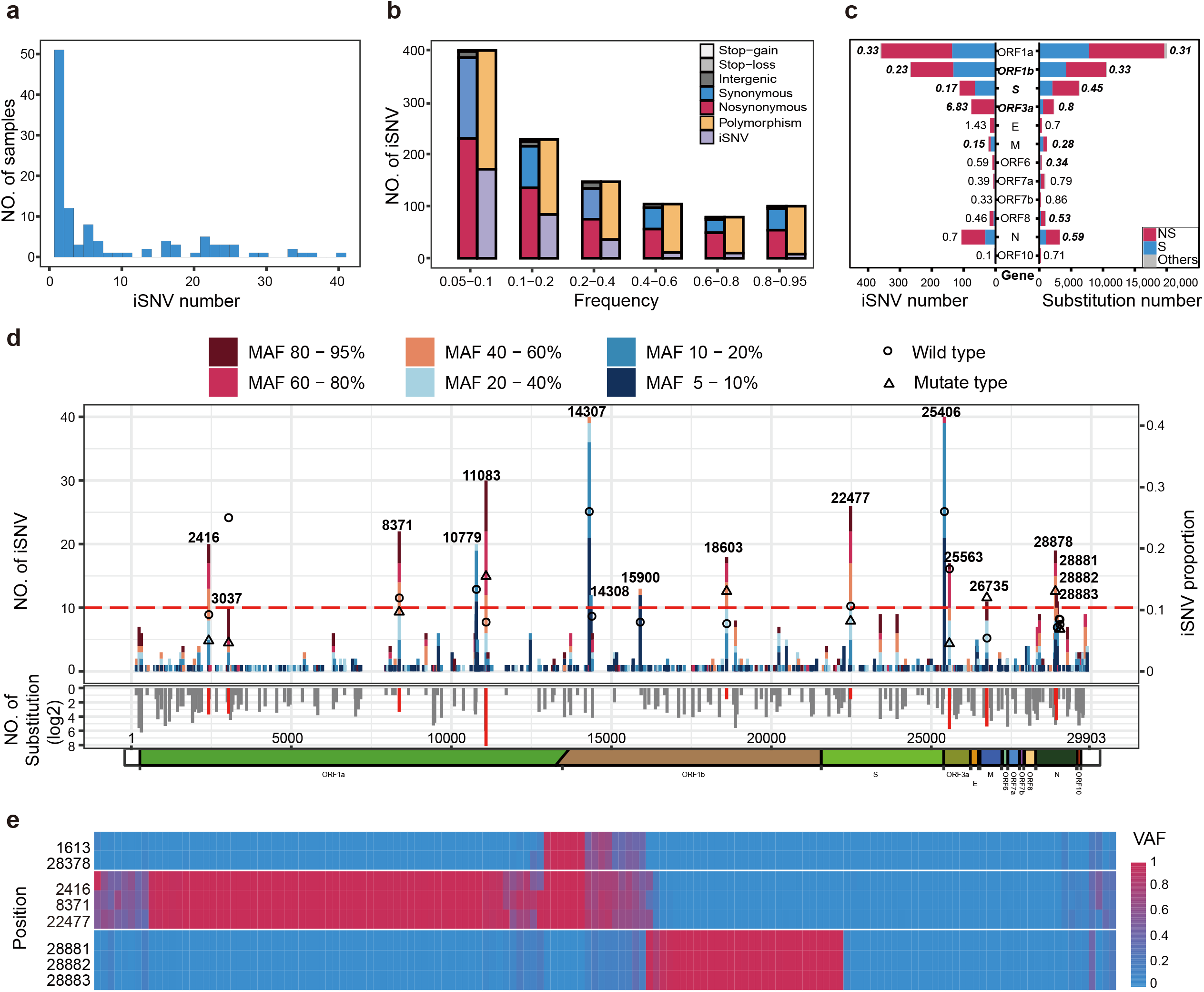
The profile of the iSNVs. **a** The distribution of the number of iSNVs per sample. **b** Mutate allele frequency distribution and mutation types of all iSNVs. The orange and the purple bar represent the number of mutations that observed and not observed in the polymorphism data. **c** The Ka/Ks ratio for each gene using the iSNVs data and the polymorphism data, the values were calculated using KaKs_Calculator2.0 software (MS model), the bold and italicized number indicates that the number was significantly different from 1, the bold and italicized gene name indicates that the Ka/Ks ratios were significantly differed between the iSNVs data and the polymorphism data. **d** The number, incidence, and genomic distribution of all iSNVs. The bar color shows the MAF of each iSNV, the circle and triangle indicate the iSNVs incidence for the wild type and the mutant type at positions with iSNV incidence no less than 10 (right y-axis, major allele frequency>=0.7, depth>=100), respectively. The red dash line represents iSNV number equals to 10. The grey histogram under the x-axis shows the substitution rate estimated from the polymorphism data. The bars at positions with iSNV incidence no less than 10 were labelled in red. **e** The concurrency of intra-host variations. Three clusters of concurrent mutations are shown, positions in each cluster tend to appear intra-host variations at the same time and no significant difference was found between their variation frequencies.

Positions with higher iSNV incidences (≥10) had higher substitution rates estimated from the polymorphism data compared to the rest of iSNV positions (Wilcoxon rank sum test, *p* < 0.01). Meanwhile, the iSNVs incidence was also positively correlated with the substitute rate (rho=0.22, *p* < 0.01, Spearman rank correlation), suggesting that a large proportion of iSNV with MAF>0.05 were neutral or mildly deleterious, which did not undergo strong purifying selection during fixation in the individual. However, two of the most mutable positions (14307, 25406) and several highly mutable positions (10779, 14308, 15900) were notable exceptions that these mutations were rarely observed in the polymorphism data and had lower MAF (Fig. 3d), probably reflecting a purifying selection or RNA editing at these positions. Of note, none of the sequence context of these mutations matched known RNA-editing motifs. Furthermore, we noted that iSNVs at position 11083 showed the highest MAF (with the mean of 0.61) and this position also had the highest substitution rate among all positions, suggesting that either the position is error-prone during replication or the mutation at this position is favored by natural selection.

Intriguingly, we found the concurrence of 7 pairs of iSNVs which involved 8 loci (r^2^ >8) in the population (Fig. 3e). Moreover, paired iSNV had similar MAF (*p* > 0.05), indicating that these mutations either occurred simultaneously in the same individual or represented a co-infection of multiple SARS-CoV-2 strains. However, only G28881A, G28882GA, and G28883C were in strong linkage disequilibrium in the polymorphism data, suggesting that other mutation pairs unlikely contributed to significant fitness benefit. However, the mechanism and consequence of these concurrent mutations were elusive and need further investigation.

### Spread of SARS-CoV-2 in Pakistan revealed one large local transmission chain

To understand the spread and transmission of SARS-CoV-2 in Pakistan, we performed both haplotype network and phylogenetic analysis (Supplementary Fig. 3) using 150 newly obtained SARS-CoV-2 genomes from Pakistan in this study and 2,135 randomly sampled high quality global public sequences (from 54,204 sequences with the collection date before June 17, and contains 14 Pakistani sequences) as of October 9, 2020. Those sequences closely related to Pakistani sequences in the network (including the parental and filial sequences of the Pakistan nodes) were further selected to infer the virus transmission chain (Fig. 4a). For all Pakistani sequences, we identified 140 distinct haplotypes (Supplementary Table 2), and most sequences can be further grouped into five distinct clusters defined as C1-C5 with 9 to 74 sequences each.

**Fig. 4.**
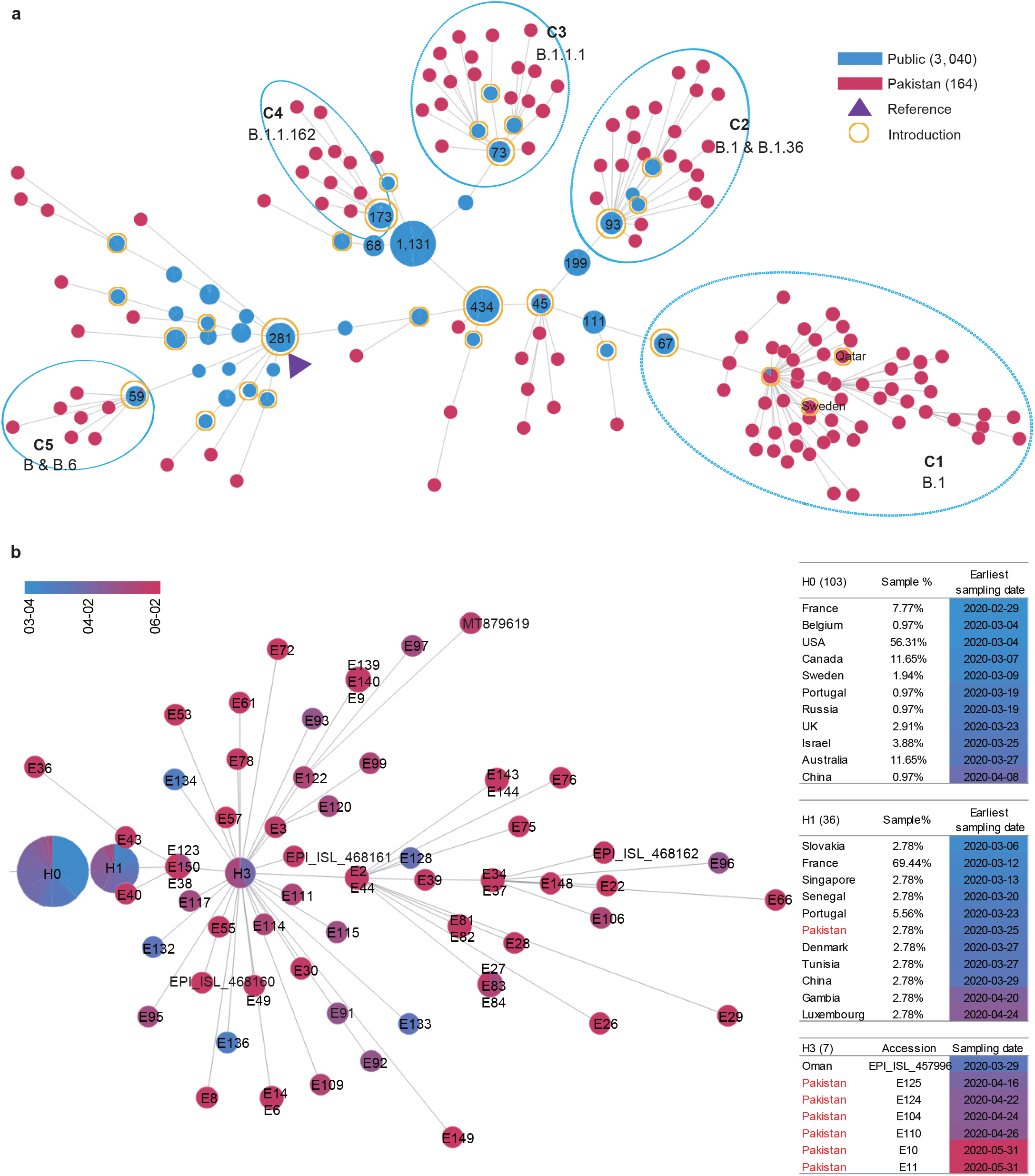
Spread and transmission of Pakistani sequences. **a** Haplotype network of all SARS-CoV-2 sequences in Pakistan (red node) and those closely-related sequences from public (blue node). Each node represents a distinctive haplotype, and the line length between any two nodes is proportional to sequences distance. Pakistani sequence clusters were labeled with C1-C5, Nodes of the reference were marked by a purple triangle, and nodes of putative introductions were labelled with yellow circle. **b** The haplotype network of Cluster 1. The color of the nodes, from blue to red, represents the sampling time from March 4 to June 2, 2020. The sample names were marked in each node. The nodes marked by H0 and H1 represent the parent and the first generation node of Pakistani sequences, respectively. The node of H2 means the super spreader sequences. The percentage of countries sampled for each node sequence is listed in the table on the right.

The largest cluster (C1) includes 74 genomes sampled from March 16 to June 2, and is characterized by a nucleotide substitution G8371T in the nonstructural proteins 3 (nsp3) region, causing an amino acid change of Q1884H (Q2707H of ORF1ab) (Table 1). Nearly all sequences in C1 could be classified as the B.1 based on Pangolin COVID-19 Lineage Assigner(19). Here, we have made a few interesting observations. First, the common ancestor sequence of C1 (node marked as H1, having the least number of variants compared to reference) consists of one Pakistani sequence (E127, collected on March 25 in Punjab) and 36 sequences from France, Slovakia, Portugal, Singapore, and Senegal (Fig. 4b). And the sampling time of most of these public sequences was earlier than that of Pakistan. By further tracing the upstream parent sequences (H0) of this Pakistani sequence, we found the sampling time of some sequences from USA, France, Portugal, and Sweden was also earlier than sample E127 and the date Pakistan began to suspend international flights (March 13, 2020), which suggested that this Pakistani sample might be early international introductions related. Secondly, another key node (marked as H3 in Fig. 4b) consists of 7 sequences, six from Pakistan and one from Oman. The haplotype network shows that the H3 node have more filial nodes, suggesting these six Pakistan cases may be super spreaders in the transmission chain. As the sampling time of Oman sequence (March 29, 2020) is earlier than those of the six Pakistani sequences (April 16 – May 31), we speculate that the Pakistan infected cases could be Oman introduction related. Thirdly, there are four descendent sequences (E132, E133, E134, and E136) that sampled earlier than Pakistani sequences in the H1 and H3 nodes. Among them, E136 collected from Federal Capital on March 16 had a travel history to Qatar. These four cases might also be imported from abroad. Fourthly, nearly all C1 sequences carried the common signature mutations (C2416T, C3037T, G8371T, C14408T, A23403G, and G25563T) which forms a specific haplotype, and those descendent sequences are mainly Pakistan specific, forming at least 2-5 transmission chains, indicating that viruses in C1 were mutated and widely transmitted in Pakistan. Moreover, most cases of C1 were collected from Punjab (30) and Federal Capital (38), two adjacent districts. The local transmission across the country could be the result of expansion of cases seeded by the local religious congregations in Lahore (the capital of Punjab) and social gatherings and later by relaxing the lock-down situation.

**Table 1.** Major clusters and signature variants of Pakistani sequences.

| Cluster | Lineage | No. of sequences<br>(this study/public) | Common variants | Putative<br>introductions<br>countries | No. of<br>putative<br>introductions | Signature<br>variants | Gene | Nuc.<br>mutation | AA<br>position<br>and<br>change |
| --- | --- | --- | --- | --- | --- | --- | --- | --- | --- |
| C1 | B.1 | 74 (70/4) | 2416(T), 3037(T),<br>8371(T), 14408(T),<br>23403(G), 25563(T) | **USA, France,**<br>**Portugal, Sweden,**<br>**Oman , Qatar** | 4 | 8371 | *orf1ab* | G>T | Q2702H |
| C2 | B.1<br>B.1.36 | 24 | 3037(T), 14408(T),<br>18877(T), 23403(G),<br>25563(T), 26735(T) | **Japan, Saudi Arabia**<br>**and Turkey. Others:**<br>India, Indonesia,<br>Kazakhstan | 3 | 26735 | *M* | C>T | - |
| C3 | B.1.1.1 | 22 (17/5) | 3037(T), 4002(T),<br>10097(A), 13536(T),<br>14408(T), 23403(G),<br>23731(T), 28881(A),<br>28882(A), 28883(C) | **UK, Others:** Austria,<br>CostaRica,<br>CzechRepublic,<br>Denmark, Israel ,<br>Netherlands, Peru ,<br>Portugal, Sweden | 4 | 4002<br>13536 | *orf1ab*<br>*orf1ab* | C>T<br>C>T | T1246I<br>- |
| C4 | B.1.1.162 | 12 | 313(T), 3037(T),<br>14408(T), 23403(G),<br>28881(A), 28882(A), | Brazil, Denmark,<br>France, Hungary,<br>India, Israel, Japan, | 1 | 313 | *orf1ab* | C>T | - |
|  |  |  | 28883(C) | Norway, Portugal,<br>Russia, Singapore,<br>SouthKorea,<br>Switzerland, United<br>Arab Emirates, United<br>Kingdom, USA |  |  |  |  |  |
| C5 | B<br>B.6 | 9 | 6312(A), 11083(T),<br>13730(T), 23929(T),<br>28311(T) | USA, Australia,<br>Canada, China,<br>Gambia, India,<br>Malaysia, New<br>Zealand, Oman,<br>Senegal, Sierra Leone,<br>Singapore | 1 | 11083 | *orf1ab* | G>T | L3606F |
|  |  |  |  |  |  | 23929 | S | C>T | - |

Note: the countries marked in bold represents the putative importing countries, and countries in italics were visited by some patients.

### Inferred multiple introductions and international transmission routes

The haplotype network analysis also revealed four clusters (C2-C5) that did not form a transmission chain which may be due to Pakistan’s local epidemic prevention and control policy.

Cluster 2 (C2), assigned as the B.1 and B.1.36 lineage, has 24 sequences which is characterized by a synonymous nucleotide substitution (C26735T) in the *orf3a* gene. Aside from this synonymous mutation, all C2 sequences carried other five mutations: C3037T, C14408T, C18877T, A23403G, and C25563T. No identical sequences to any of the ones in C2 were found in public databases. To find the possible source of introduction of viruses in C2, the upstream parent node (H0) of C2 Pakistani sequences were looked into. There were 93 sequences sampled from six Asian countries, including India, Indonesia, Japan, Turkey, Kazakhstan, and Saudi Arabia (Fig. 5a), and three derived nodes consisting of 13 sequences collected from Malaysia, Saudi Arabia, Turkey, India, and the Netherlands. More importantly, the sampling time of 46 sequences from Japan, Saudi Arabia, and Turkey was earlier than that of Pakistani sequences in C2 and the flight closing date of Pakistan. We therefore inferred that there were most probably three independent international introductions to Pakistan from Japan, Saudi Arabia, and Turkey.

**Fig. 5.**
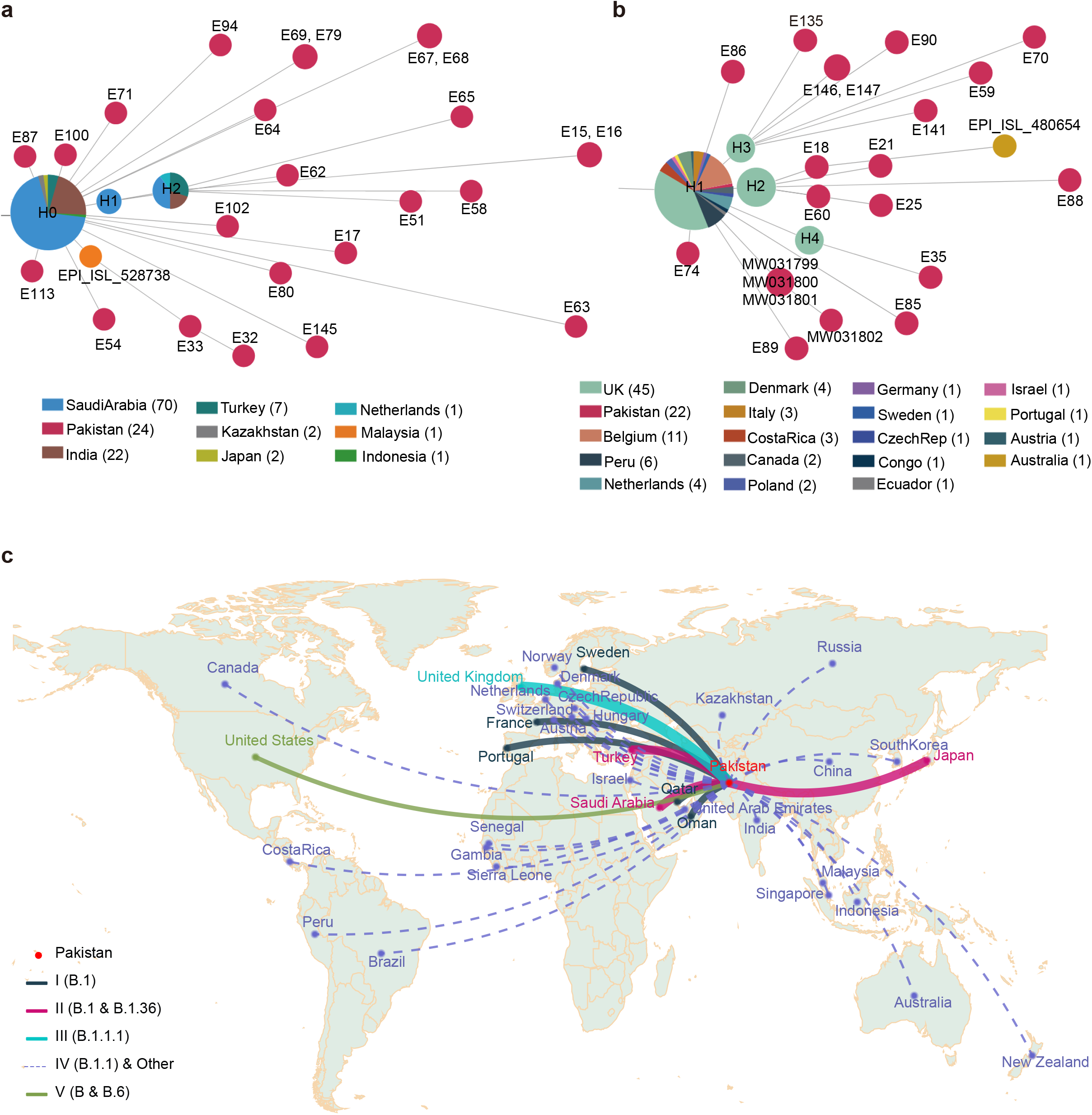
Two representative of introduction-related clusters and schematic diagram of inferred international introduction routes. **a-b** Haplotype network of cluster 2 and cluster 3. **c** Schematic diagram of inferred global imports and introductions. The solid lines represent those putative importing countries and the dotted lines represent those likely importing countries. The thickness of the lines is proportional to the number of inferred introductions.

There are 22 Pakistani sequences (18 generated from this study and 4 collected from public databases) in Cluster 3 (C3), which were designed as the B.1.1.1 lineage(19). C3 is characterized by two nucleotide substitutions (C4002T and C13536T) in the *orf1ab* gene, and the mutation of C4002T causes an amino acid change T1246I. Thirteen sequences in C3 were directly connected to sequences from United Kingdom, while the rest 9 were associated with those spreading in countries including Austria, Costa Rica, Czech Republic, Denmark, Israel, Netherlands, Peru, Portugal, and Sweden (Fig. 5b). We speculate that there were at least four international introductions according to the haplotype network.

Cluster 4 (C4) contains 10 sequences and is designated as the B.1.162 lineage. When tracing the parent node of C4 sequences, we found 173 sequences distributed in nearly 20 countries, such as Denmark and France, which suggested the source of C4 cases in Pakistan could be complex. From the haplotype network distribution, we speculate that there were about one or more independent international introduction in C4 (Fig. 4a).

For Cluster 5 (C5), there are 9 sequences that can be traced back to a sequence sampled on March 20 in the USA. The descendant sequences spreading in more than 10 countries by adding two additional mutation sites (G11083T and C23929T), among which 2 are Pakistani samples (E108 and E118) collected in the middle of April. So, we inferred that they might be introduced from the United States, Australia, Canada, Guangdong/Taiwan, Gambia, India, Malaysia, New Zealand, Oman, Senegal, Sierra Leone, or Singapore.

In all, by tracing the parent node of all clusters using haplotype network method, we inferred several putative introduction routes (solid line in Fig. 5c), including United Kingdom, France, Portugal, Sweden, Japan, Saudi Arabia, and Turkey, and multiple likely introductions (dashed lines in Fig. 5c). Based on the number of network links, we inferred that there were about 28 introductions. To further confirm the likelihood of early introductions, we compared the beginning time of epidemic between Pakistan and neighboring countries and those putative or likely importing countries (Supplementary Fig. 4), and found that several countries had outbreaks earlier than Pakistan, except for Saudi Arabia and Oman.

## DISCUSSION

This is the largest genomic epidemiology study of COVID-19 in Pakistan, with 150 high quality complete SARS-CoV-2 genomes obtained and analyzed. SARS-CoV-2 genome evolves at a rate of approximately 1-2 mutations per month(25). Hundreds of mutation sites and 1057 iSNV were identified, some of which were found over-represented in this cohort sequences, suggestive of widely spreading in Pakistan. High sequence diversity was also reflected by 136 different haplotypes among 150 sequenced genomes. On the one hand, this may be due to a long sampling time span, and most of the samples were collected in the peak of outbreak. On the other hand, it may indicate that there are continuous importations of different virus types.

An unprecedented sequencing depth was obtained in this study for SARS-CoV-2 genome due to the implementation of an efficient hybridization enrichment method, which enables not only the detection of the mutations, but also the analysis of the iSNVs. The iSNV could represent either in vivo evolution of virus after infection or infection of different virus strains simultaneously or successively. Although we identified several mutations that tended to occur concurrently and showed similar frequencies, they cannot be explained by coinfection of a secondary strain that was observed in our dataset. We suspect that these concurrent mutations may be explained by strong compensatory effects of epistasis(26,27). Thus, these positions in strong linkage disequilibrium would be of particularly interest to researchers working on functionality of SARS-CoV-2 mutations. The long tail distribution of the number of iSNVs in each sample agree with several recent findings(28,29), and the fast-evolution of the virus in some patients may result in the generation of new strains with excess number of mutations(30,31). Most iSNVs occurred at postions with high substitute rate, suggesting a large fraction of mutations were neutral despite that SARS-CoV-2 genome was overall under purifying selection(29,32). In contrast, we noted that two of the most frequently observed iSNVs in our data were located at positions that were not polymorphic at the population level, suggesting that they were either dysfunctional mutations subject to intensive purifying selection or caused by in vivo modification, such as RNA editing. However, we noted that they were not located in known context recognized by APOBEC or ADAR gene families. Besides, our results emphasized that some individual-shared mutations were indeed iSNVs, which might arise independently after infection, how they impact the performance of genome epidemiology merit further investigation.

The mutations in Pakistan could be used to infer the source of the virus and the transmission chain of SARS-CoV-2 within the country. Furthermore, such data can also be helpful to monitor the export of virus to other countries. Based on the phylogenetic and haplotype network analysis, we inferred about 28 independent introductions during the three-month study period. Notably, viruses in one dense cluster C1 had been widely transmitted in the community, spanning KPK and Punjab regions. The fast spread of the epidemic may be due to a mass religious gathering in Lahore (the capital of Punjab) held during March 11-12, 2020. It has been reported that COVID-19 transmissions were often associated with clusters of cases linked to gatherings, including ones in work-places(33), churches(34), and especially in close living environment such as care homes and homeless(35). These social gathering based viral clusters are thought to often involve superspreading(35). Nearly all C1 sequences (73/74) were directly or indirectly linked to several Pakistani sequences in the H1 and H3 nodes, and formed a transmission chain of up to five generations. To better control the second wave of the epidemic in Pakistan, any social gatherings whatsoever, should be strictly controlled. On the other hand, the fast transmission ability of C1 might have been contributed by its characteristic mutation G8371T in the nsp3 protein. To test this possibility, we collected all public sequences with the G8371T mutation and built a haplotype network (Supplementary Fig. 5a). Several local clusters spreading with multiple generations were found, suggesting that this mutation may enhance viral transmission. The *nsp3* gene encodes a predicated phosphoesterase which is a component of the replication–transcription complexes (RTCs), and the interactions of N and nsp3 are essential that links the viral genome for processing. Some residues involved in interactions between nsp3 and C-terminal domain (CTD) of N protein have been predicated which could be a potential drug target for future inhibitors(36). Moreover, 188 mutations (1∼9 per isolate) occurred after the introduction of the two virus isolates in Pakistan. Whether these mutations alone or in combination could increase the transmissibility of the virus warrants further investigation.

Based on our haplotype network tracing analysis, the early epidemic in Pakistan was related to those international introductions, as the parent nodes of all clusters and those sporadic individuals are directly connected with viruses outside Pakistan. For example, the parent node sequences of C1 containing C241T/C3037T/C14408T/A23403G/G25563T, was reported to be associated with an international business conference in Boston resulting in extensive international spread and low-level community transmission in Europe(37). Their C2416T/G8371T sub-lineage was likely the first in the C1 cluster imported into Pakistan based on the tMRCA (the most recent common ancestor) estimation - the median estimated time to tMRCA of the C1 cluster was March 10, 2020 (with 95% CI: March 7-14, 2020)(25). The mutation appeared frequently among SARS-CoV-2 genomes from Europe and USA in public databases (81 genomes as of Feb. 19, 2021), suggesting its introductions from USA or Europe. Genomes of another sister cluster C2, harboring the C26735T variant in addition to common signatures of C241T/C3037T/C14408T/A23403G/C18877T/G25563T, have been found mainly in Asia (93 genomes as of Oct. 9, 2020). The other two clusters, C3 and C4, were derivatives from European lineages B.1.1.1 and B.1.1.162, respectively. Although Pakistan closed its land borders and limited international flights in the middle of March, viruses directly related to the ones outside the country have continued to be isolated in Pakistan. This revealed that viruses might be imported into Pakistan through other routes, such as cold-chain transportation in the frozen food industry(38). It is therefore important for Pakistan to strengthen the inspection and quarantine of imported cold-chain food and pay more attention to the personal protection of relevant workers to better prevent and control COVID-19. Notably, based on globally haplotype network analysis (Supplementary Fig. 5b), almost no progenies of any of the 150 genomes in this study can be found out of Pakistan, indicating an effective response and control measures to prevent the virus spreading out of Pakistan.

In summary, we have generated a total of 150 high quality and complete SARS-CoV-2 genomes of Pakistan, and identified 29 over-represented mutations. The genomic epidemiology revealed five distinctive spreading clusters. Among them, C1 was the largest cluster and introduced putatively from USA, France, Portugal, Sweden, Oman, and Qatar. The deep hierarchical structure of C1 indicated an extensive and persistent nation-wide transmission of the virus which is probably contributed by a signature mutation G8371T in the *nsp3* gene. No progenies of any of these viruses have been found outside of Pakistan, most likely due to the nonphmarcological intervention to control the virus spread. Besides, more than twenty putative or likely international introductions were inferred from Europe and the Middle East, and several of them were consistent with the result of epidemiological investigation.

## Supporting information

Supplementary Figures, and will be used for the link to the file on the preprint site.

## Data Availability

The raw sequence data and the whole genome sequences reported in this paper have been deposited in the Genome Sequence Archive under accession number CRA003122 and Genome Warehouse under accession number GWHAOJE01000000~GWHAOOX01000000, in National Genomics Data Center, Beijing Institute of Genomics (China National Center for Bioinformation), Chinese Academy of Sciences.

https://bigd.big.ac.cn/gsa/browse/CRA003122

https://ngdc.cncb.ac.cn/search/?dbId=gwh&q=PRJCA003179&page=1

## DATA AVAILABILITY

The raw sequence data and the whole genome sequences reported in this paper have been deposited in the Genome Sequence Archive(39) and Genome Warehouse(40) in National Genomics Data Center(41), Beijing Institute of Genomics (China National Center for Bioinformation), Chinese Academy of Sciences, under accession number CRA003122 that are publicly accessible at https://bigd.big.ac.cn/gsa, and GWHAOJE01000000-GWHAOOX01000000 at https://bigd.big.ac.cn/gwh. The whole genome sequences also have been deposited in GenBank(42) under accession number MW421982-MW422131 at https://www.ncbi.nlm.nih.gov/genbank/.

## ACKNOWLEDGEMENT

Complete genome sequences used for analyses were obtained from the CNCB-NGDC, CNGBdb, GenBank, GISAID, and NMDC databases. We acknowledge the sample providers and data submitters listed in Supplementary Table 4.

## AUTHOR CONTRIBUTIONS

S. Song were responsible for conceptualization, methodology, and manuscript writing. C. Li, L. Kang, D. Tian, W. Ma, and S. Zhao analyzed the sequence data. N. Badar was in charge of sample collection along with A. Ikram, M. Salman, and M. Umair. A. Abbasi, H. Shireen, and F. Batool executed metadata collection. X. Jiang, C. Wang, B. Zhang, Y. Sun, W. Li, M. Lei, Sh. Li, and Q. Qi performed sequencing library construction. H. Chen and Y. Yang supervised the study. M. Li, Y. Xue and Y. Bao conceived experiments and supervised the study, reviewed and edited manuscript. All authors read and approved the final manuscript.

## FIGURE LEGEND

**Supplementary Fig. 1 Epidemic in Pakistan**. Distribution of daily confirmed cases, deaths, and detection numbers in Pakistan as of Oct 9, 2020. The dates marked in red represent the first reported cases, Pakistan announced upgrading the prevention and control, and announced the closure of all international flights, respectively.

**Supplementary Fig. 2 Sequencing coverage and genomic variants in Pakistan SARS-CoV-2 genomes. a** Distribution of sequencing depth, percentage of reference sequenced covered, and number of bases covered above 30X. **b** Correlations between SARS-CoV-2 read counts, genome coverage and Ct value. The number of SARS-CoV-2 reads for all samples in the unit of reads per million (RPM), proportion of the genome with sequencing depth≧1 and ≧100, and the Ct value below the detection limit (39) was replaced by 42 for better visualization. Spearman’s rank correlation coefficient and *p* value are shown in the figure. **c** Distribution of sequence number for different number of variants.

**Supplementary Fig. 3 Haplotype network and phylogenetic tree of 150 viruses sampled from Pakistan (colored in red) on a background of 2**,**135 randomly sampled high quality global public sequences (from 54**,**204 sequences with the collection date before June 17 2020. a** Haplotype network. **b** Phylogenetic tree.

**Supplementary Fig. 4 Comparison the epidemic development between Pakistan and neighboring countries and those putative importing countries**.

**Supplementary Fig. 5 Evolution and transmission analysis as of Feb 22, 2021. a** Haplotype network of all sequences with 8371 mutation. **b** Haplotype network of sequences that conneted with Parkistanis.

## TABLE LEGEND

**Supplementary Table 1** Summary of population density, GDP income, Multidimensional poverty index, and COVID-19 outcomes in major provinces of Pakistan.

**Supplementary Table 2** Meta information, sequence variants, lineage and cluster classification for Pakistani samples.

**Supplementary Table 3** Variants identified in SARS-CoV-2 genome sequences of Pakistan.

**Supplementary Table 4** All meta information for public SARS-CoV-2 sequences used in this study.

