## Supplementary Figures, and will be used for the link to the file on the preprint site. for "Genomic epidemiology of SARS-CoV-2 in Pakistan"

### Supplementary Fig.1

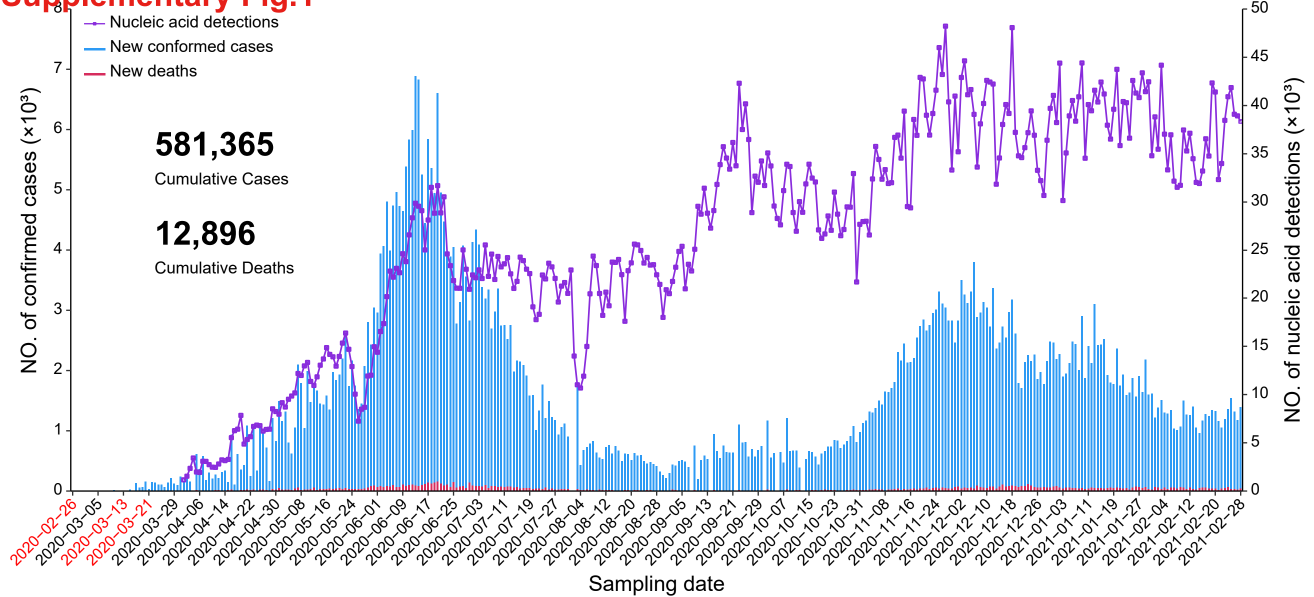

Supplementary Fig.2

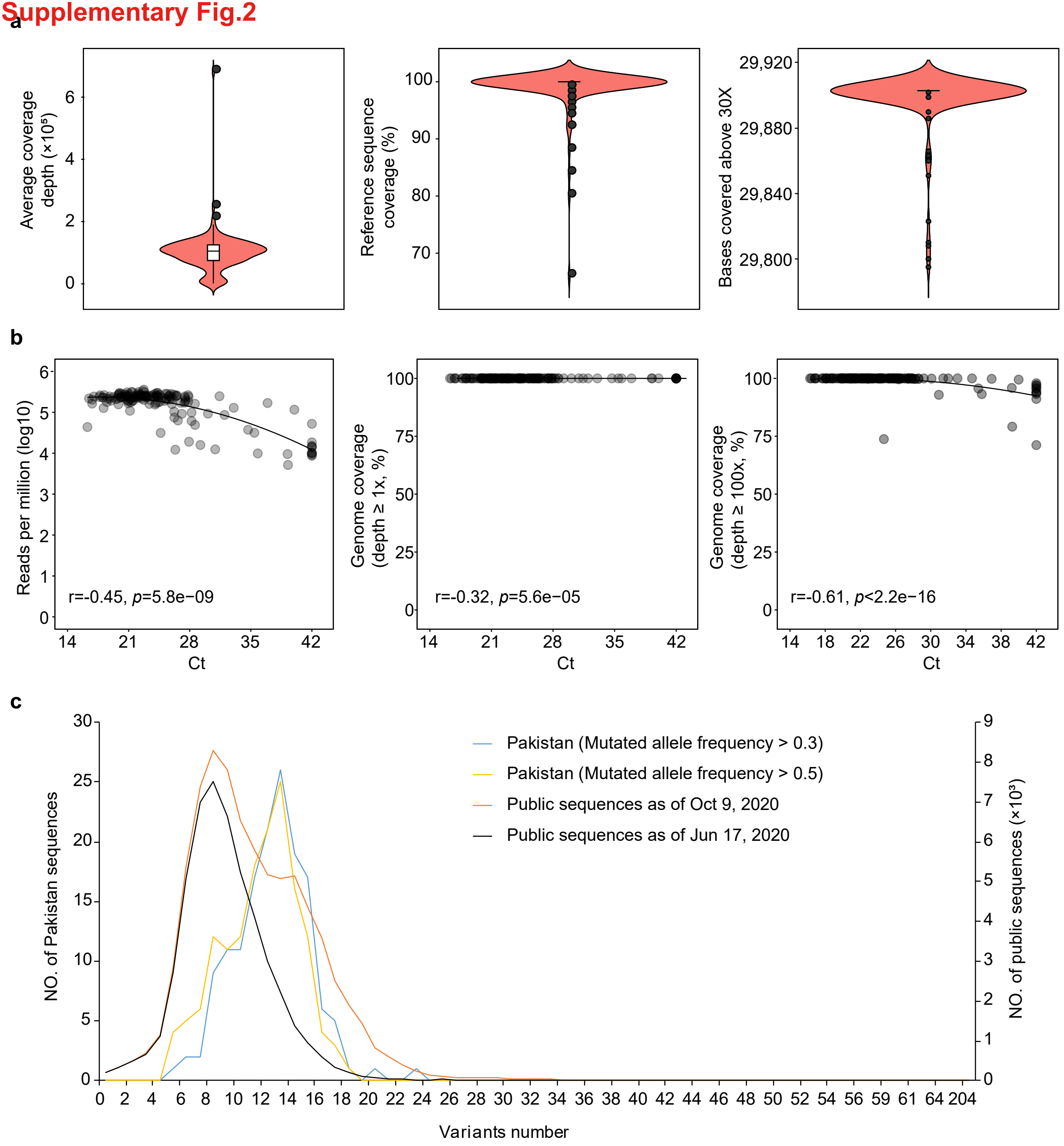

#### Supplementary Fig.3

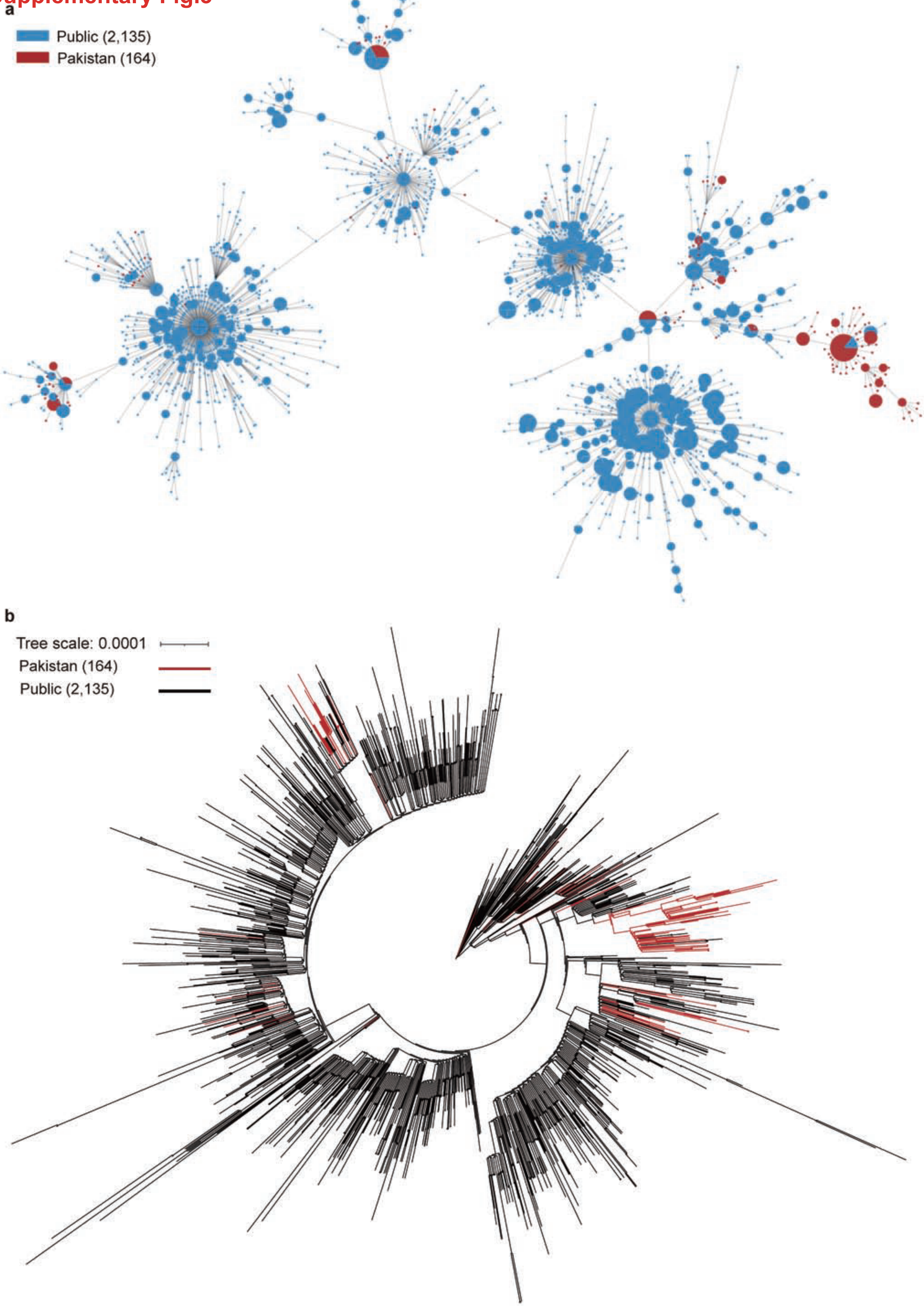

### Supplementary Fig.4

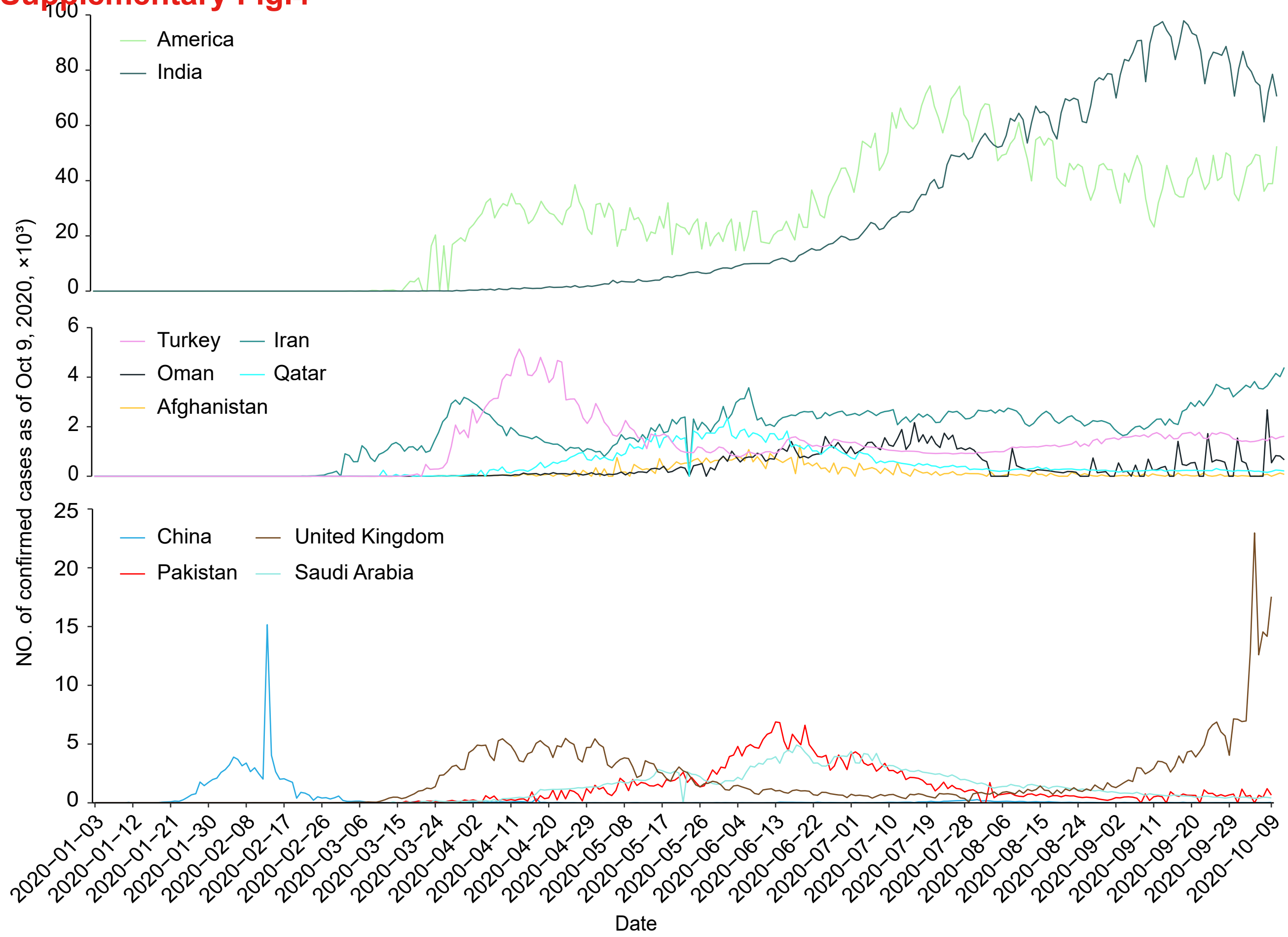

Supplementary Fig.5

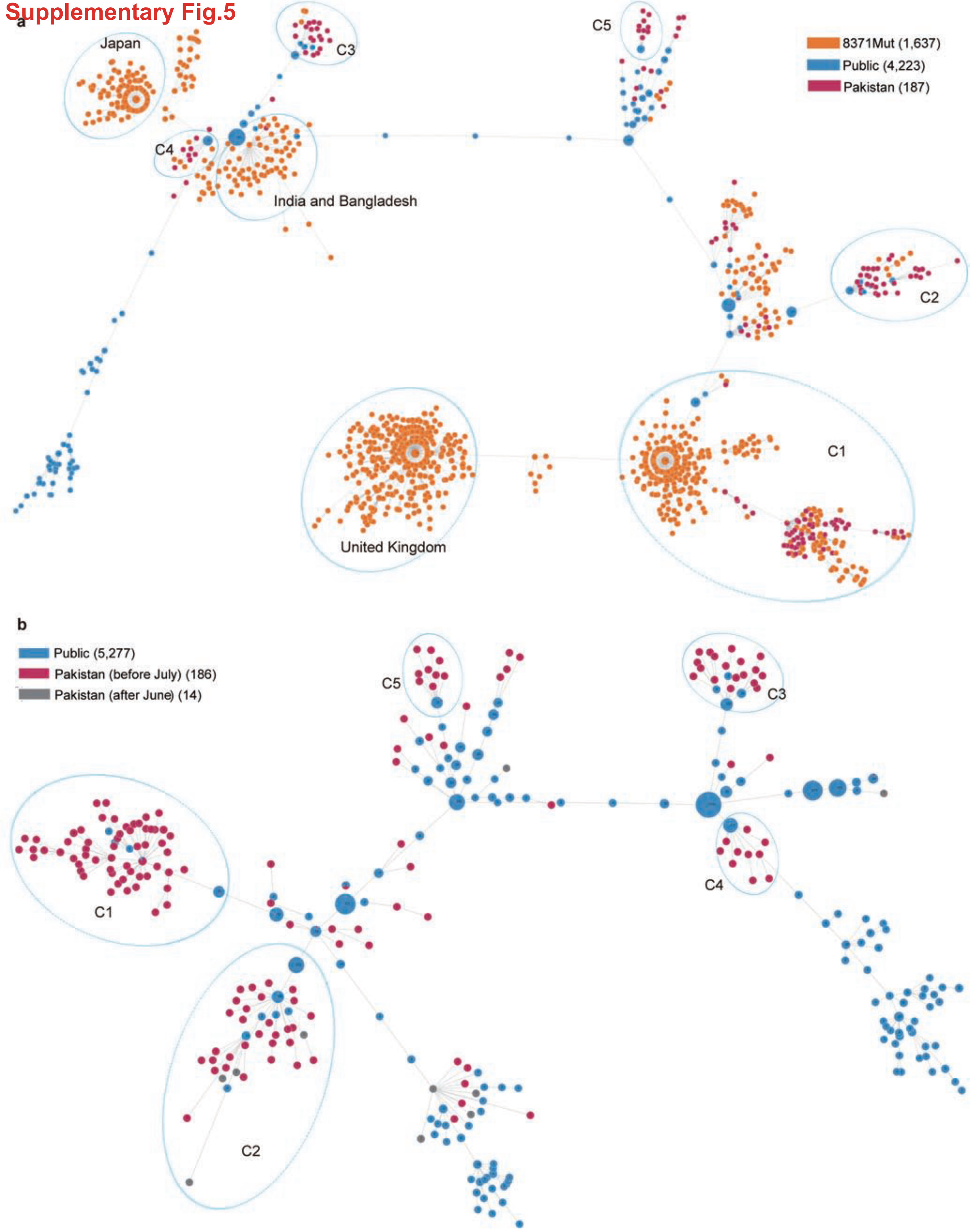
